# Aging and Mortality among People with Diagnosed HIV in Italy: Recent Trends and Projections

**DOI:** 10.64898/2026.02.02.26345395

**Authors:** Alex Viguerie, Vincenza Regine, Lucia Pugliese, Barbara Suligoi

**Affiliations:** Department of Pure and Applied Sciences, Università degli studi di Urbino Carlo Bo. 60129 Urbino, Italy; National AIDS Unit, Department of Infectious Diseases, Italian National Institute of Health, 00161 Rome, Italy

**Keywords:** HIV/AIDS, aging populations, demographic trends, mathematical modeling, age-structured modeling

## Abstract

The advent of antiretroviral therapy (ART) has led to substantial increases in life expectancy among persons with diagnosed HIV (PWDH), and in turn, an increasingly older population. This represents a public health challenge, as older PWDH are more susceptible to age-related health morbidities compared to the general population. In this study, we triangulate diverse data sources to reconstruct the Italian PWDH age structure over the past decade to better-understand recent trends, and provide demographic projections through 2035.

We find that the PWDH population grew from approximately 112,000 persons in 2012, to 140,000 in 2024, and forecasted to reach 155,000 by 2035. This is primarily driven by decreased PWDH mortality, with such decreases forecast to continue. Persons over 60, estimated as 8.6% of the PWDH population in 2012, had increased to over 20% by 2020, and are projected to reach 47.2% by 2035. By 2030, over 10% of PWDH in Italy are projected to be over 75, compared to less than 1% in 2012.

Our results demonstrate that the Italian HIV care infrastructure must prepare for a dramatic shift from managing a predominantly young-adult disease to caring for a majority-elderly population within the next decade, representing an unprecedented transformation in the nature and scope of required services.

## Introduction

Since the advent of effective antiretroviral therapies, the life expectancy of people diagnosed with HIV (PWDH) has increased significantly. In developed countries, it is now common for PWDH to live up to 70 years and beyond [1], [2], [3]. Although this is undoubtedly a positive development, it poses new challenges for health systems. PWDH present age-related chronic conditions more frequently and at a younger age compared to the general population; contributing factors include the specific effects of HIV infection in older adults, the long-term effects of the infection, and prolonged use of antiretroviral therapy (ART) [1], [4], [5], [6]. Increasingly, therefore, HIV prevention and care interventions must include the management of health complications associated with HIV in the elderly, in long-term infections, and in prolonged ART use among PWDH.

To this end, effective methods for accurately predicting age-related chronic conditions among PWDH are fundamental for resource allocation, long-term strategy planning, and informing possible interventions. Having reliable methods to clearly quantify and predict the future age structure of the PWDH population is a necessary step toward this goal. However, this is not a trivial task. As mortality trends in the PWDH population differ from those of the general population, the population size relatively small, and the data often noisy, many techniques commonly used in general population demographic analysis are unsuitable for PWDH populations [2], [3], [7], [8], [9], [10], [11].

In [3], the authors have recently developed a multi-stage computational pipeline that combines ensemble demographic reconstruction based on partial differential equations (PDEs), data assimilation, and scientific machine learning techniques to provide reconstructions and forecasts for the PWDH population in the United States. In the present work, we apply the same techniques to the Italian PWDH population.

## Methods

We used age-structured mortality data among PWDH with a previous AIDS diagnosis (2012-22) and new diagnosis data (2012-23), provided by the Istituto Superiore di Sanità [12]. Since age-structured HIV prevalence data were not available, we triangulated several sources, specifically [13], [14], [15], together with surveillance data in subsequent years, to estimate age-specific HIV prevalence in 2011.

Starting from this initial point, we then updated the population structure using the provided diagnosis and mortality data. The rest of the computational procedure described in [3], including ensemble Kalman inversion to estimate mortality rates, ensemble population sampling for uncertainty quantification, and non-negative dynamic mode decomposition (nnDMD) to predict the evolution of mortality rates and HIV diagnoses, was employed with minimal modification.

We remark that, in contrast to the US data, substantial changes in PWDH mortality trends during the COVID-19 pandemic were not observed in the Italian data, and therefore no adjustments were necessary.

To account for uncertainty in the data, we independently sampled 1,000 initial population distributions from our reconstructed 2011 PWDH demographic distribution. Each ensemble member was simulated over the entire period; the results reported herein are the ensemble means. Full credible ranges for our projections, obtained using the ensemble variance, are provided in the full supplemental data.

### Modeling and computational aspects

We will now discuss the core components of our modeling and computational pipeline in further detail. Given the aim of the present work, we focus on the components of the model that directly affect identifiability and inference. As such, the interested reader is encouraged to consult [3] for a more comprehensive and mathematically detailed description.

### Demographic evolution and mortality reconstruction

Let *u*(*a, t*), *μ*(*a, t*), *λ*(*a, t*) denote, respectively the total number, probability of mortality, and new HIV diagnoses among PWDH aged *a* at time *t*. The age-structure of population evolves from *t* to *t* + 1as:

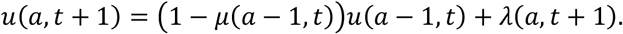

To determine *μ*, which is unknown, we use the above relation, together with surveillance data, which report annual PWDH deaths over discrete five-year age ranges [*a*_*k*_, *a*_*k*+4_]:

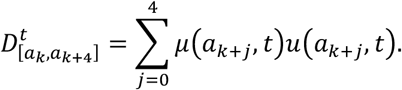

Note the above relation is not identifiable directly – we must recover the value of *μ*at five distinct ages from only *one* data point 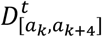. To this end, we introduce additional statistical structure by considering an *ensemble* of populations *u*_*i*_(*a, t*). The overall demographic structure of the *u*_*i*_ are consistent at initialization-that is, the number of PWDH in each five-year age band is the same for each *u*_*i*_. However, *within* each age band, considerable variability exists between the *u*_*i*_. Similarly, we also define a separate *μ*_*i*_ for each ensemble population *u*_*i*_ as well as separately-sampled *λ*_*i*_. Finally, we will make use of the shorthand notation:

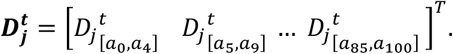

By introducing the additional ensemble structure, we may identify the mortality hazard *μ*as a statistical estimator ensuring optimality over the entirety of the ensemble space. Specifically, we apply the inverse ensemble Kalman filtering algorithm (InvEnKF) to achieve this.

The algorithm is briefly described as follows. Assuming *m* total ensemble elements, for each year *t*, we apply the algorithm below for until convergence. The *k ™*th iteration reads:

1. Advance the population states *u*_*i*_with the demographic equation.
2. Compute:

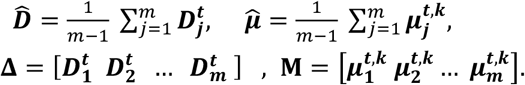
3. Compute:

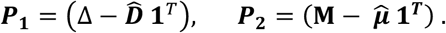
4. Update estimate of ***μ***:

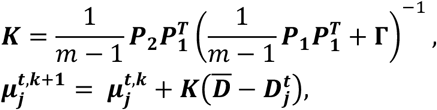 Where 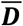 is the vector containing the surveillance deaths in each age interval and **Γ** is a diagonal matrix accounting for noise inherent in the data.
5. Check convergence. If met, advance to time ***t*** + 1. Otherwise, repeat.

Interested readers may refer to [16] for a more detailed discussion of the InvEnKF algorithm.

### Non-negative dynamic mode decomposition

Non-negative dynamic mode decomposition (nnDMD) is a data-driven forecasting technique introduced in [3] for settings where the number of temporal observations is low, and the data are inherently nonnegative (as is the case with mortality hazards). nnDMD is a variant of dynamic mode decomposition (DMD); we refer the reader to e.g. [17], [18], [19], [20] for more details.

Briefly, nnDMD works as follows. Let *μ*_*t*_ be a d-dimensional vector whose entries contain age-specific mortality hazards in year *t*. We then consider a sequence of mortality hazards over *n* years: *μ*_1_, *μ*_2_, …, *μ*_*n*_ and arrange them into two *d ×*(*n* − 1) matrices:

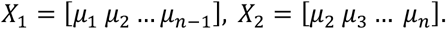

nnDMD is then recovered as the solution to the following constrained least-squares problem:

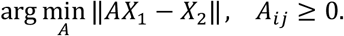

In other words, nnDMD finds the matrix *A* such that

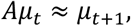

while guaranteeing that the *μ*remain nonnegative. A more in-depth discussion, including empirical validation and mathematical analysis for problems of this type, can be found in [3].

## Results

Our results show that the PWDH population in Italy has aged significantly in the last decade. An immediate effect is the increase in the PWDH population in Italy, which has grown from ∼112,000 in 2012 to ∼140,000 in 2024. We predict a continuation of this growth, which should reach ∼150,000 around 2030, and approach 155,000 by 2035 (Figure 1, Table 1). This growth is not due to an expected increase in transmission and diagnoses; in fact, we predict a decline in new diagnoses (Table 2). Instead, the increase in population is mainly caused by the continued decrease in mortality among PWDH, particularly those at younger ages, resulting in a greater number of PWDH reaching advanced ages (Figures 2-4, Tables 1-3).

**Table 1:** Simulated PWDH prevalence, 2024-35.

| Age | 2024 | 2025 | 2026 | 2027 | 2028 | 2029 | 2030 | 2031 | 2032 | 2033 | 2034 | 2035 |
| --- | --- | --- | --- | --- | --- | --- | --- | --- | --- | --- | --- | --- |
| 0-15 | 1 | 1 | 1 | 1 | 1 | 1 | 1 | 1 | 1 | 1 | 1 | 1 |
| 15-20 | 51 | 57 | 63 | 68 | 72 | 73 | 73 | 71 | 69 | 66 | 63 | 60 |
| 20-25 | 436 | 437 | 460 | 489 | 515 | 532 | 537 | 533 | 521 | 505 | 486 | 466 |
| 25-30 | 2061 | 1847 | 1688 | 1589 | 1554 | 1556 | 1563 | 1563 | 1555 | 1536 | 1506 | 1465 |
| 30-35 | 5567 | 5001 | 4530 | 4132 | 3799 | 3534 | 3315 | 3124 | 2975 | 2879 | 2820 | 2767 |
| 35-40 | 10618 | 9855 | 9045 | 8295 | 7643 | 7039 | 6467 | 5956 | 5507 | 5118 | 4794 | 4513 |
| 40-45 | 13020 | 13184 | 13289 | 13157 | 12726 | 12061 | 11259 | 10410 | 9612 | 8902 | 8245 | 7623 |
| 45-50 | 18636 | 17086 | 15707 | 14764 | 14369 | 14374 | 14518 | 14575 | 14387 | 13905 | 13189 | 12326 |
| 50-55 | 22348 | 22577 | 22575 | 22138 | 21139 | 19718 | 18159 | 16759 | 15781 | 15333 | 15284 | 15371 |
| 55-60 | 20578 | 21304 | 21859 | 22263 | 22587 | 22866 | 23061 | 23033 | 22572 | 21567 | 20148 | 18575 |
| 60-65 | 17859 | 18153 | 18637 | 19300 | 20043 | 20772 | 21422 | 21943 | 22335 | 22647 | 22914 | 23132 |
| 65-70 | 15452 | 16674 | 17347 | 17641 | 17774 | 17910 | 18176 | 18644 | 19289 | 20015 | 20731 | 21349 |
| 70-75 | 6622 | 7979 | 9739 | 11696 | 13616 | 15273 | 16461 | 17128 | 17424 | 17557 | 17695 | 17938 |
| 75-80 | 3405 | 3883 | 4387 | 4928 | 5575 | 6481 | 7791 | 9485 | 11377 | 13237 | 14840 | 16002 |
| 80-85 | 1631 | 1838 | 2092 | 2429 | 2840 | 3283 | 3742 | 4223 | 4743 | 5371 | 6253 | 7546 |
| 85-100 | 1448 | 1684 | 1939 | 2209 | 2504 | 2839 | 3215 | 3646 | 4164 | 4772 | 5444 | 6170 |
| TOT | 139733 | 141560 | 143358 | 145099 | 146757 | 148312 | 149760 | 151094 | 152312 | 153411 | 154413 | 155304 |

**Table 2:** Simulated deaths among PWDH, 2024-35.

| Age | 2024 | 2025 | 2026 | 2027 | 2028 | 2029 | 2030 | 2031 | 2032 | 2033 | 2034 | 2035 |
| --- | --- | --- | --- | --- | --- | --- | --- | --- | --- | --- | --- | --- |
| 0-15 | 0 | 0 | 0 | 0 | 0 | 0 | 0 | 0 | 0 | 0 | 0 | 0 |
| 15-20 | 0 | 0 | 0 | 0 | 0 | 0 | 0 | 0 | 0 | 0 | 0 | 0 |
| 20-25 | 1 | 1 | 1 | 1 | 1 | 1 | 1 | 1 | 1 | 1 | 1 | 0 |
| 25-30 | 1 | 1 | 1 | 1 | 1 | 1 | 1 | 1 | 1 | 1 | 1 | 1 |
| 30-35 | 4 | 3 | 3 | 2 | 2 | 2 | 2 | 2 | 1 | 1 | 1 | 1 |
| 35-40 | 11 | 9 | 8 | 7 | 6 | 6 | 5 | 5 | 4 | 4 | 3 | 3 |
| 40-45 | 12 | 12 | 10 | 11 | 10 | 9 | 8 | 8 | 7 | 6 | 5 | 5 |
| 45-50 | 31 | 29 | 24 | 22 | 20 | 19 | 19 | 19 | 18 | 17 | 16 | 15 |
| 50-55 | 68 | 68 | 65 | 62 | 59 | 54 | 49 | 44 | 39 | 36 | 35 | 34 |
| 55-60 | 116 | 118 | 118 | 117 | 114 | 112 | 109 | 106 | 102 | 95 | 87 | 78 |
| 60-65 | 70 | 66 | 69 | 69 | 70 | 70 | 70 | 69 | 69 | 67 | 66 | 65 |
| 65-70 | 56 | 70 | 69 | 71 | 69 | 68 | 66 | 65 | 65 | 65 | 66 | 66 |
| 70-75 | 42 | 49 | 55 | 65 | 75 | 83 | 89 | 92 | 92 | 90 | 88 | 86 |
| 75-80 | 36 | 39 | 41 | 46 | 49 | 55 | 63 | 74 | 87 | 100 | 110 | 117 |
| 80-85 | 15 | 17 | 18 | 21 | 23 | 26 | 29 | 32 | 34 | 38 | 42 | 49 |
| 85-100 | 16 | 21 | 23 | 27 | 30 | 33 | 37 | 41 | 46 | 50 | 55 | 60 |
| TOT | 479 | 503 | 505 | 522 | 529 | 539 | 548 | 559 | 566 | 571 | 576 | 580 |

**Figure 1.**
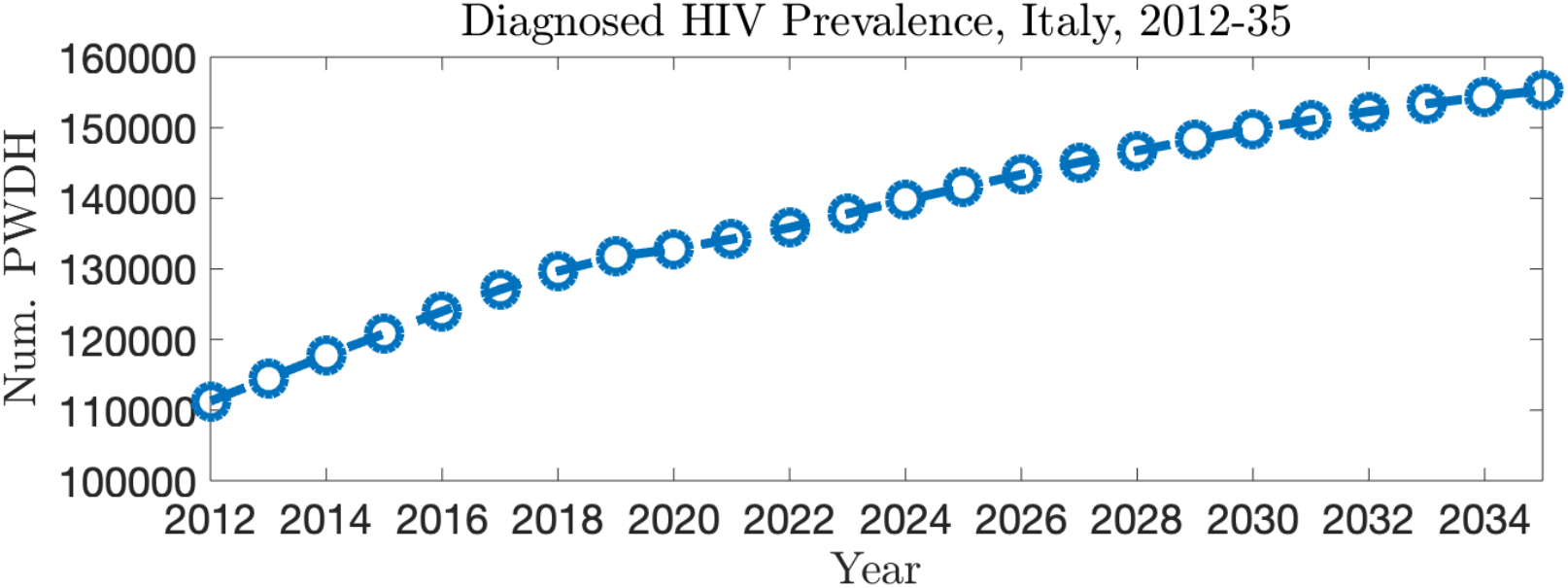
Total PWDH population in Italy, 2012-35.

**Figure 2.**
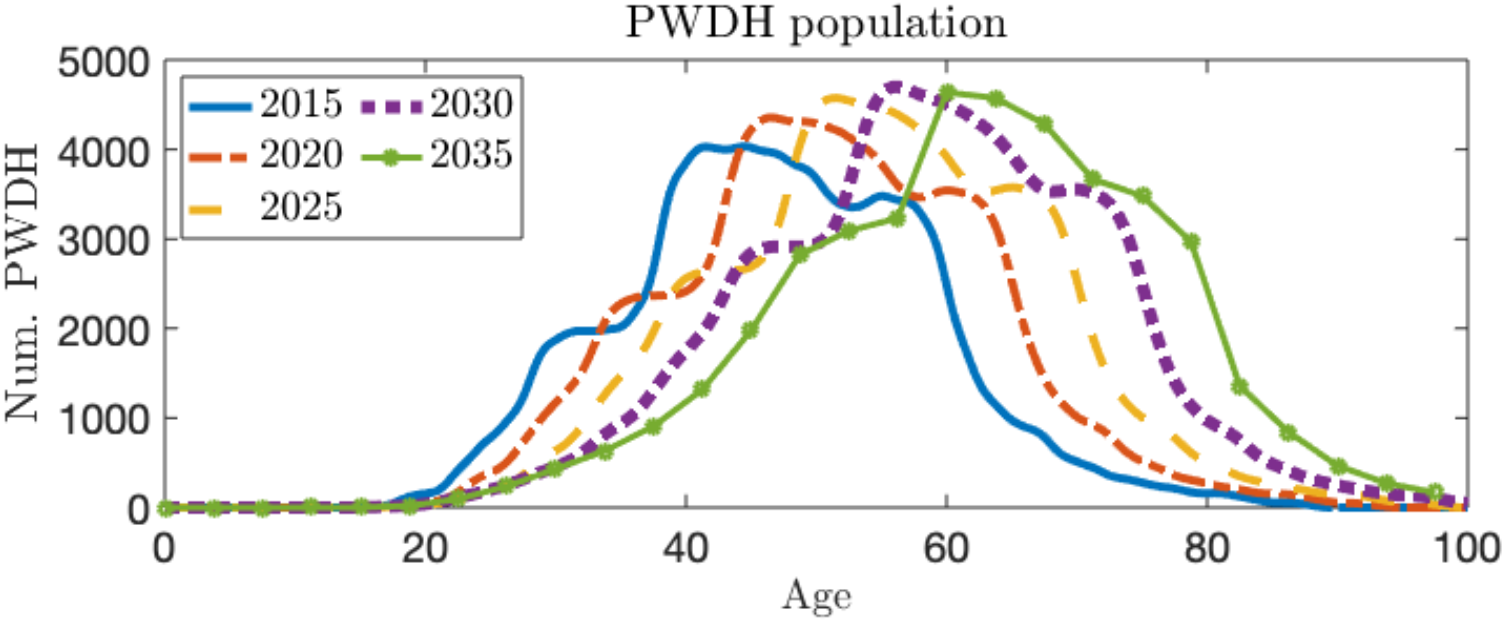
Demographic profile of the Italian PWDH population: evolution in time and forecast progression

Going into more detail, in line with [13], we estimate that people aged 60 or older represented 8.6% of PWDH in 2012 and 11.0% in 2014. By 2020, we estimate that this share increased to 21.8%, and we predict that 47.2% of Italian PWDH will be over 60 by 2030. The effect is even more marked in the over-75 age group: we estimate that only 1% of PWDH were over 75 in 2012, while the share rose to almost 5% in 2024 and should reach 10% by 2030 (Figures 2 and 3, Table 1).

**Figure 3.**
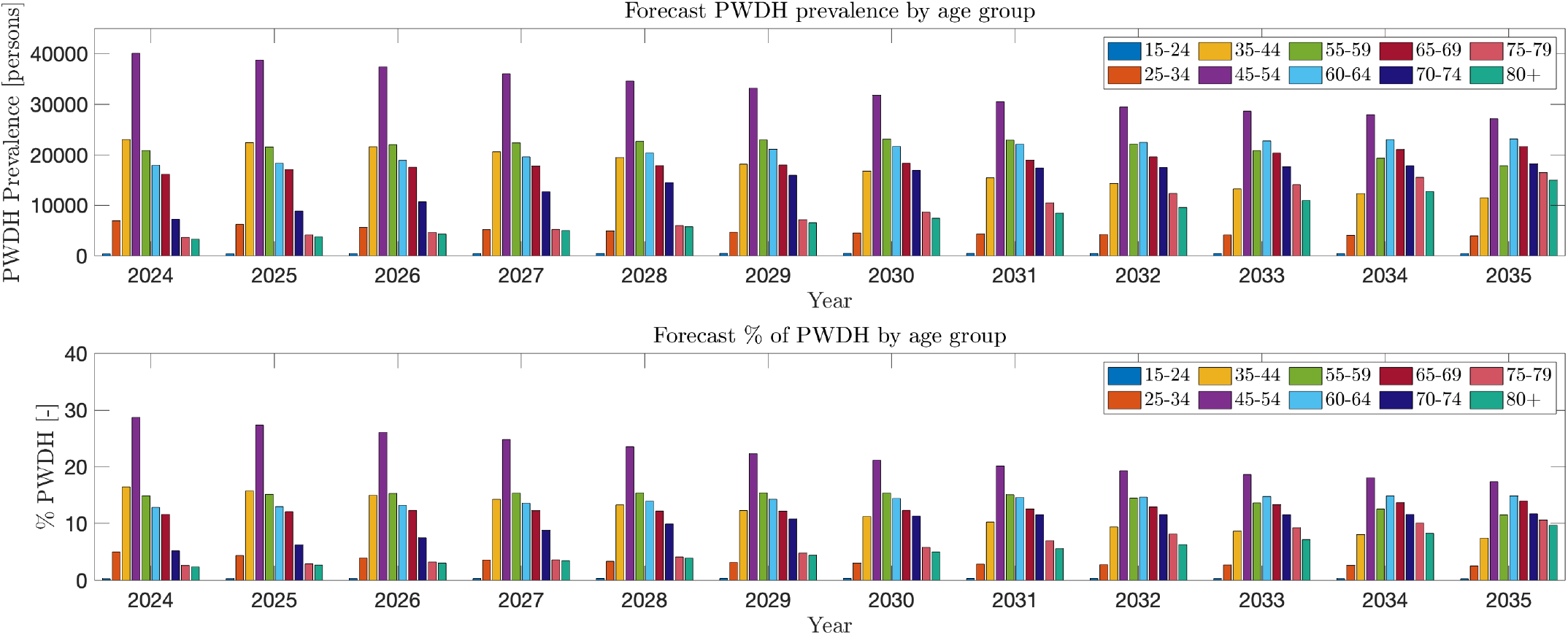
Demographic profile of PWDH in Italy, temporal evolution in discrete age groups, 2024–35

This change is determined by a shift in mortality trends among PWDH with a previous AIDS diagnosis. Among this cohort, we predict a further decline in mortality, especially at younger ages, in continuity with long-term trends. Consequently, the percentage of PWDH deaths occurring in the oldest age groups is destined to increase significantly. Deaths among PWDH aged 75 or older represented less than 3% of PWDH deaths in 2012 and have steadily increased in recent years. We predict the continuation of this trend: deaths in this group will exceed 20% of PWDH deaths by 2029, 30% by 2032, and 35% by 2035 (Table 2, Figure 4).

**Figure 4.**
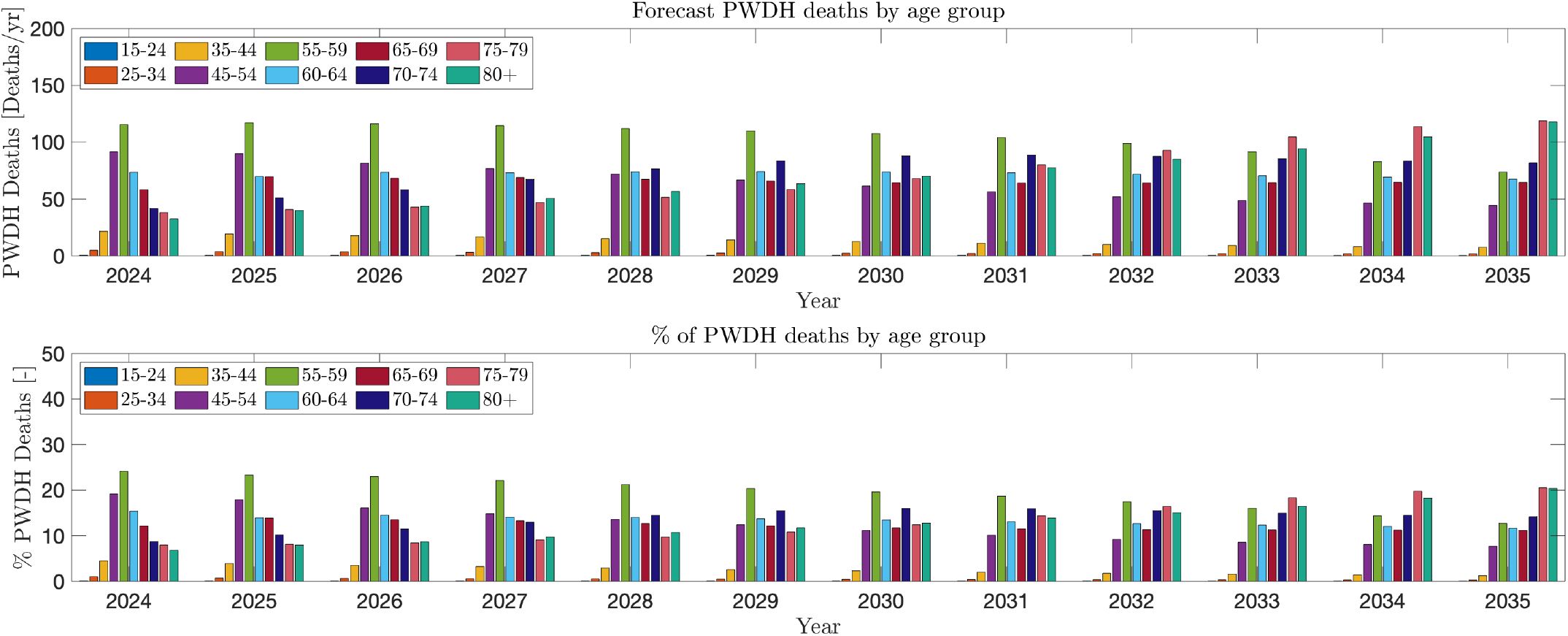
Deaths among PWDH in Italy by age group, 2024–35

Regarding new HIV diagnoses, we project continued, steady decrease in the future, with annual new diagnoses dropping below 2000 after 2030. We forecast declines across all age groups, with little changes in the relative age distribution of new HIV diagnoses, in line with pre-existing trends (Table 3). These trends contribute to a skewed age profile, with pronounced asymmetry around the modal age, reflecting the accumulation of long-surviving cohorts at older ages and a smoother, more extended distribution toward younger ages, particularly visible by the end of the forecast period (Figure 2, Table 1).

**Table 3:** Simulated new HIV diagnoses, 2024-35.

| Age | 2024 | 2025 | 2026 | 2027 | 2028 | 2029 | 2030 | 2031 | 2032 | 2033 | 2034 | 2035 |
| --- | --- | --- | --- | --- | --- | --- | --- | --- | --- | --- | --- | --- |
| 0-15 | 2 | 2 | 2 | 2 | 2 | 2 | 2 | 2 | 2 | 2 | 2 | 1 |
| 15-20 | 26 | 30 | 34 | 36 | 36 | 35 | 34 | 32 | 31 | 29 | 28 | 26 |
| 20-25 | 121 | 148 | 165 | 171 | 170 | 165 | 159 | 151 | 144 | 137 | 131 | 124 |
| 25-30 | 272 | 263 | 290 | 292 | 288 | 278 | 266 | 253 | 242 | 230 | 219 | 209 |
| 30-35 | 346 | 286 | 316 | 312 | 305 | 293 | 280 | 268 | 255 | 243 | 232 | 221 |
| 35-40 | 305 | 314 | 306 | 302 | 293 | 282 | 271 | 260 | 248 | 237 | 226 | 216 |
| 40-45 | 305 | 281 | 300 | 297 | 289 | 278 | 266 | 254 | 242 | 231 | 220 | 209 |
| 45-50 | 264 | 290 | 294 | 288 | 276 | 262 | 249 | 237 | 226 | 215 | 205 | 195 |
| 50-55 | 247 | 249 | 231 | 228 | 221 | 212 | 202 | 193 | 184 | 175 | 166 | 158 |
| 55-60 | 189 | 223 | 171 | 155 | 150 | 145 | 139 | 133 | 127 | 121 | 115 | 109 |
| 60-65 | 138 | 118 | 94 | 88 | 85 | 81 | 78 | 74 | 71 | 67 | 64 | 61 |
| 65-70 | 88 | 54 | 52 | 51 | 50 | 48 | 46 | 44 | 42 | 40 | 38 | 36 |
| 70-75 | 47 | 35 | 32 | 29 | 27 | 26 | 25 | 24 | 23 | 22 | 20 | 20 |
| 75-80 | 20 | 16 | 14 | 12 | 11 | 10 | 10 | 9 | 9 | 9 | 8 | 8 |
| 80-85 | 4 | 6 | 5 | 4 | 4 | 3 | 3 | 3 | 3 | 3 | 3 | 3 |
| 85-100 | 2 | 1 | 1 | 1 | 1 | 1 | 1 | 1 | 1 | 1 | 1 | 1 |
| TOT | 2376 | 2316 | 2307 | 2268 | 2208 | 2121 | 2031 | 1938 | 1850 | 1762 | 1678 | 1597 |

## Discussion

Our study is subject to several important limitations that should be considered when interpreting these results. In Italy, deaths among PWDH are recorded only among PWDH with a previous AIDS diagnosis, a group that has represented approximately 25% of new diagnoses in recent years [12], reflecting long-standing patterns of testing and diagnosis. Consequently, mortality data unavailable for PDWH without AIDS may lead to overestimation of the number of people living with HIV and disproportionately reflect outcomes among individuals with advanced disease at diagnosis, potentially biasing estimates of age-specific mortality. However, because the majority of older PWDH acquired HIV infection many years ago, we expect the qualitative patterns identified here—particularly those related to population ageing and the shifting distribution of deaths toward older age groups—to remain largely reliable. This is especially true over the short-to-medium term (approximately 5–10 years), which is the primary time horizon relevant for health system planning.

Our projections of future HIV diagnoses and PWDH mortality trends are based on a fully data-driven approach. While this strategy avoids reliance on explicit structural assumptions— particularly regarding mortality, where general population life-table methods are poorly suited to the PWDH population—it necessarily assumes continuity in the underlying processes shaping recent trends. In this sense, causal drivers of diagnoses and mortality are incorporated implicitly through historical data. Rapid or substantial changes in HIV prevention, testing, or care in the coming years could therefore affect the accuracy of our forecasts. Examples include large-scale changes in PrEP uptake, shifts in HIV testing strategies, improvements in early diagnosis, developments in ART efficacy, or shifts in long-term ART adherence (for example, due to increased use of long-acting injectables [21]). Should such changes occur, future outcomes may diverge from the patterns observed in the historical record, particularly beyond the short-term projection window.

Despite these limitations, our findings provide a clear and consistent picture of the ongoing demographic transformation of the PWDH population in Italy. The substantial growth observed over the past decade, and projected to continue in the coming years, is driven primarily by declining mortality and increasing survival rather than by increases in HIV transmission or new diagnoses. As a result, an increasing proportion of PWDH are surviving into older age groups, leading to a rapid expansion of the population aged 60 years and older, and particularly those aged 75 years and above.

This demographic shift has important implications for public health planning and HIV care delivery. An ageing PWDH population is likely to experience a higher burden of chronic comorbidities, polypharmacy, and functional decline, requiring closer integration between HIV services, multidisciplinary health approaches, geriatric care, and management of non-communicable diseases. Surveillance systems and care models that were originally designed for a younger population of PWDH may need to be adapted to reflect the growing importance of long-term care, multimorbidity, and age-related health needs [2], [22]. Anticipating these changes through demographic forecasting is therefore essential for informing resource allocation, workforce training, and the design of future HIV prevention and care strategies in Italy.

## Data Availability

All data produced in the present study are available upon reasonable request to the authors.

## Declarations

The authors declare that no funds, grants, or other support were received during the preparation of this manuscript.

The authors have no relevant financial or non-financial interests to disclose.

All authors contributed to study design and conception. Surveillance data were provided by B. Suligoi, V. Regine, and L. Pugliese. Data analysis, software implementation, and tabulation were performed by A. Viguerie. Original draft was written by A. Viguerie. All authors contributed to revision/editing of final manuscript.

